# The potential impact of Omicron and future variants of concern on SARS-CoV-2 transmission dynamics and public health burden: a modelling study

**DOI:** 10.1101/2021.12.12.21267673

**Authors:** Epke A Le Rutte, Andrew J Shattock, Nakul Chitnis, Sherrie L Kelly, Melissa A Penny

## Abstract

SARS-CoV-2 variant Omicron (B.1.1.529) was classified as a variant of concern (VOC) on November 26, 2021. (1, 2) The infectivity, severity, and immune evasion properties of Omicron relative to the Delta variant would determine 1) the probability of dominant future transmission, and 2) the impact on disease burden. (3, 4) Here we apply individual-based transmission model OpenCOVID to identify thresholds for Omicron’s or any VOC’s potential future dominance, impact on health, and risk to health systems; and identify for which combinations of viral properties, current interventions would be sufficient to control transmission. We show that, with first-generation SARS-CoV-2 vaccines (5) and limited physical distancing in place, the threshold for Omicron’s future dominance was primarily be driven by its degree of infectivity. However, we identified that a VOC’s potential dominance will not necessarily lead to increased public health burden. Expanded vaccination, that includes a third-dose for adults and child vaccination strategies, was projected to have the biggest public health benefit for a highly infective, highly severe VOC with low immune evasion capacity. However, a highly immune evading variant that becomes dominant would likely require alternative measures for control, such as strengthened physical distancing measures, novel treatments, and second-generation vaccines. These findings provide quantitative guidance to decision-makers at a critical time while Omicron’s properties are being assessed and preparedness for new VOC’s is eminent. (6) We emphasize the importance of both genomic and population epidemiological surveillance.

## Introduction

SARS-CoV-2 has been mutating continuously since its emergence in December 2019, leading to viral variants with varying infectivity, severity, and immune evading properties. On 26 November 2021, the World Health Organization identified Omicron (B.1.1.529) as a new variant of concern (VOC) after approximately seven months of Delta variant (B.1.617.2) dominating global transmission. (1) Within one week, 24 countries had reported cases of the Omicron variant, (7) with infections also occurring in previously infected and double-vaccinated individuals. (8) By mid-January 2022, more than 50% of all sequenced cases in Europe were caused by Omicron. (9) We include BA.2 in Omicron’s figures as it is a sub-lineage of B.1.1.529. Whilst scientific research assessing the infectivity, severity, and immune evasion properties of Omicron has been ongoing, understanding the potential scope of the associated public health burden of Omicron and future variants of concern (VOC) is of top priority. (2)

In many European settings, high vaccination rates - particularly among those most at risk of severe disease - have led to a reduction in disease burden. However, protection of the most vulnerable and maintaining low levels of SARS-CoV-2 transmission in the northern hemisphere has been under threat at the time of Omicron emergence for multiple reasons. Firstly, indoor contacts have been increasing during the cooler season. Secondly, immunity of the most vulnerable population (people 65+ and those with comorbidities) had started to wane since they were primarily vaccinated in the first quarter of 2021. (10) Thirdly, relaxation of most non-pharmaceutical interventions (NPIs) since the summer period of 2021 and fatigue of COVID-19 restrictions had led to increased contacts. (11) The emergence of Omicron in December 2021 combined with these threats called into question whether the vaccination strategies identified at the time would be sufficient.

Mathematical models have been used to represent transmission dynamics of SARS-CoV-2 and have supported decision-makers throughout the pandemic on the implementation and relaxation of control strategies. (12-16) Here we further developed and applied OpenCOVID, an individual-based model of SARS-CoV-2 transmission and COVID-19 disease, which includes seasonality patterns, waning immunity profiles, vaccination and NPI strategies, and properties for multiple variants. (15) We applied the OpenCOVID model to represent a general European setting and simulated the emergence of a novel variant. We analysed disease dynamics for a wide range of infectivity, severity, and immune evasion properties – relative to the Delta VOC – representing a large range of potential Omicron or any other VOC properties. This allowed us to determine the potential for Omicron to become the new dominant variant, and to estimate its potential future public health burden. We further identified combinations of properties for which first-generation vaccines, in the absence of strong NPIs, would be sufficient to contain transmission and public health burden (i.e., new SARS-CoV-2 infections, hospital occupancy, and COVID-19 related deaths) and identified those combinations for which additional measures would be required.

## Results

### Disease dynamics projections

We explored a wide range of potential infectivity and severity levels (between zero and two) of the new Omicron variant relative to Delta, and the full range of immune evading capacity from 0% (similar to Delta) to 100% (no protection from current vaccines and previously acquired natural immunity after infection). Disease dynamics were simulated over a six-month period beginning 1 December 2021 when Omicron emerged, and the cooler weather arrived. We assume an initial effective reproduction number of 1.2, the absence of strict NPIs (e.g., lockdowns), and vaccination coverage by risk group as described in Supplementary Table 1.

We compared two future vaccination scenarios. Firstly, expanded vaccination, with the administration of a third-dose six months after the second dose for individuals older than 12 years of age, and administration of vaccines for children aged 5-11 years. Secondly, we simulated a scenario with no future vaccine rollout (no third-dose vaccination in over 12-year olds and no vaccination of 5-11 year-olds). The latter leading to decay of population-level vaccine-induced immunity over time.

### Drivers of Omicron’s potential dominance

Infectivity and immune evasion were identified as the drivers behind Omicron’s potential dominance, with negligible effect from increasing severity. For an infectivity factor of 1, a 25% immune evasion property was predicted sufficient for dominance within six months, while for a 100% immune evading property an infectivity factor of 0.5 (half the infectivity of Delta) was predicted sufficient. All combinations of properties are presented in Supplementary Figures 1-2. When highly immune evading, Omicron’s chance to dominate would even slightly increase with future expanded vaccination, as vaccines provide a high level of protection against Delta variant infection (17), consequently increasing the relative susceptibility of the population to Omicron infection.

### Disease burden

If Omicron or any other new VOC would have been less infective than Delta (below 1), with no immune evading capacity, it would likely not become dominant and the public health impact due to its emergence would be negligible (Supplementary Figures 2 -3 and Supplementary Table 2). Assuming Omicron or any other new VOC would have had a partial immune evading capacity of 20% and be 20% more infectious than Delta, we predicted it would have taken approximately 4 months for it to become dominant. Although becoming the dominant variant, we predicted that these properties would lead to a relatively low increase in public health burden, particularly with expanded vaccination (as is illustrated by the white area to the right of the red curve in the left panels of Fig. 1). However, in scenarios where Omicron, or any other new variant, would have had higher levels of immune evasion capacity (50%), an increased infectivity of 50%, and be equally or more severe than Delta, dominance would occur within 5 weeks, resulting in a higher projected public health burden.

**Fig. 1:**
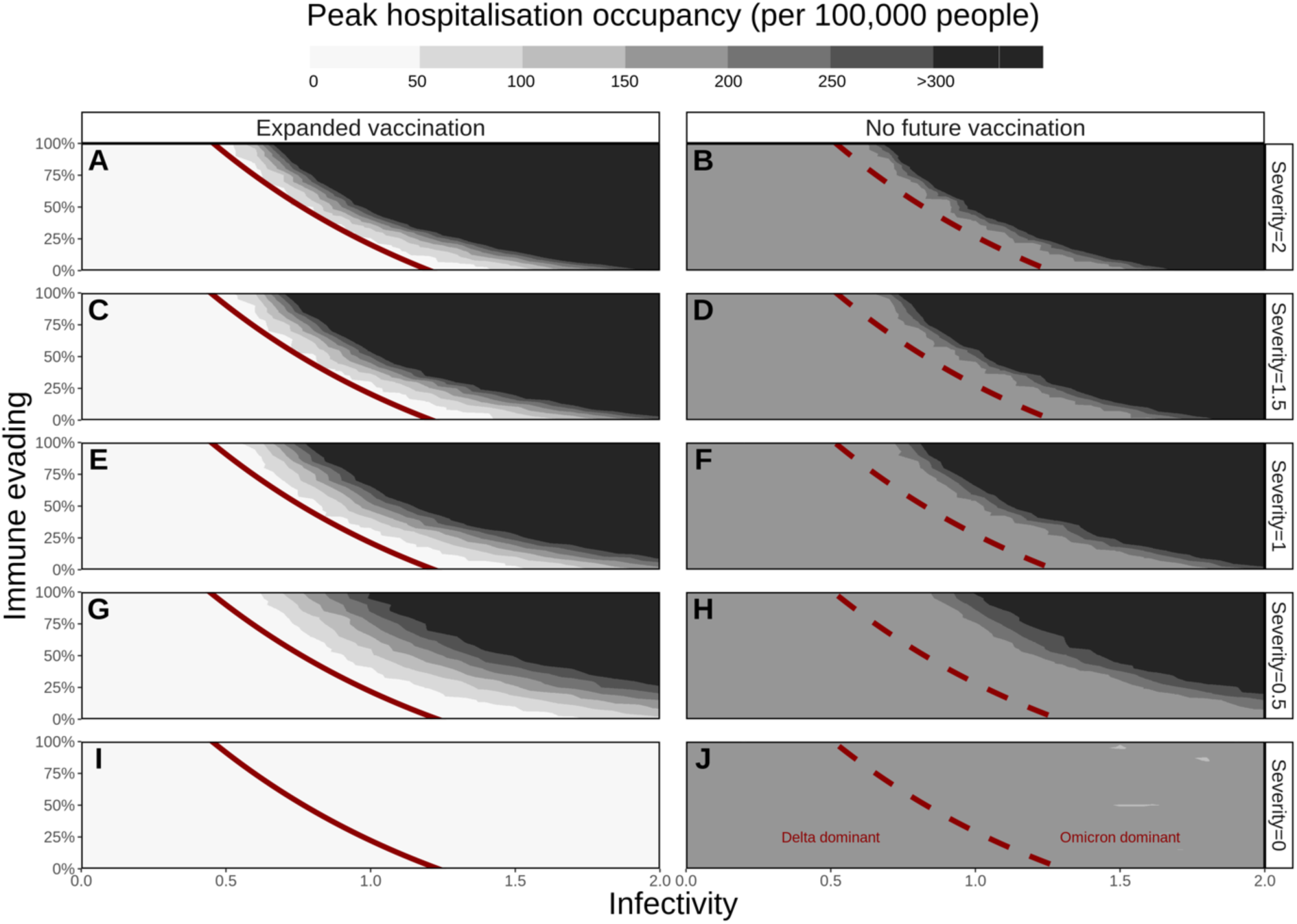
Peak daily hospital occupancy (number of beds per 100,000 population over the six-month simulation period) for a range of variant properties. The threshold (50% prevalence, see Supplementary Figure 3) for Omicron’s dominance is presented by red lines (solid for expanded vaccination, dashed for no future vaccination). Expanded vaccination includes third-doses for adults (six-months after second-dose) and vaccinating 5-11-year-olds. Rows represent Omicron’s potential severity (0 to 2) relative to Delta (1). Horizontal axes represent the range of Omicron’s potential infectivity (0 to 2) relative to Delta (1), left vertical axes Omicron’s potential immune evading capacity (0 to 100%). The peak hospital occupancy per 100,000 people has been capped at 300 per 100,000, a value that ranges for European countries between 220 (Sweden) and 800 (Germany). (18)

### Projected impact on peak hospital occupancy

Expanded vaccination was predicted to be sufficient to prevent high hospital occupancy (dark grey/black area of Fig. 1, capped at 300 hospital beds per 100,000 people) for settings in which Delta would remain dominant. (18) For a highly infectious (infectivity factor >1.5) but non-immune-evading variant, we predicted that expanded vaccination would be sufficient to prevent high hospital occupancy, however this probability diminished with increasing severity. Moreover, we explored a range of infectivity and immune evading properties for which Omicron would become dominant, but was not expected to lead to high peak hospital occupancy under the assumption of expanded vaccination (white area on the right side of the red curve in Fig. 1 and dotted area in Fig. 3). Similarly, this range diminished with increasing severity. For a variant with infectivity and severity similar to Delta, we predicted that expanded vaccination would be insufficient to prevent high peak hospital occupancy for an immune evading property above 75%. For properties that are predicted to lead to high hospital occupancy, settings would benefit from additional measures such as physical distancing, or new treatments and next generation vaccines as they become available.

### Impact on COVID-19 infections and deaths

Omicron’s potential range of severity levels was found to have negligible influence on new infections; however, it would heavily influence future mortality (in the absence of additional measures that would likely be implemented before such high mortalities (and hospital occupancy) would be reached, (Supplementary Figures 4 and 5). Fig. 2 presents the percentage of infections and deaths averted through expanded vaccinations for all specified combinations of properties. The highest public health burden in terms of infections and deaths (top right corner of Supplementary Figures 4 and 5), would be least likely to be reduced by expanded vaccination (top right of panels in Fig. 2), because of highly immune evading capacity. However, with Omicron becoming the dominant variant, expanded vaccination was predicted to have an impact on deaths for combinations of properties where severity was lower than Delta (for severity of 0.5 and 0.0 (Fig. 2). The benefit of expanding vaccination will also be seen for scenarios where Delta remains dominant (area to the left of the black lines in Fig. 2).

**Fig. 2:**
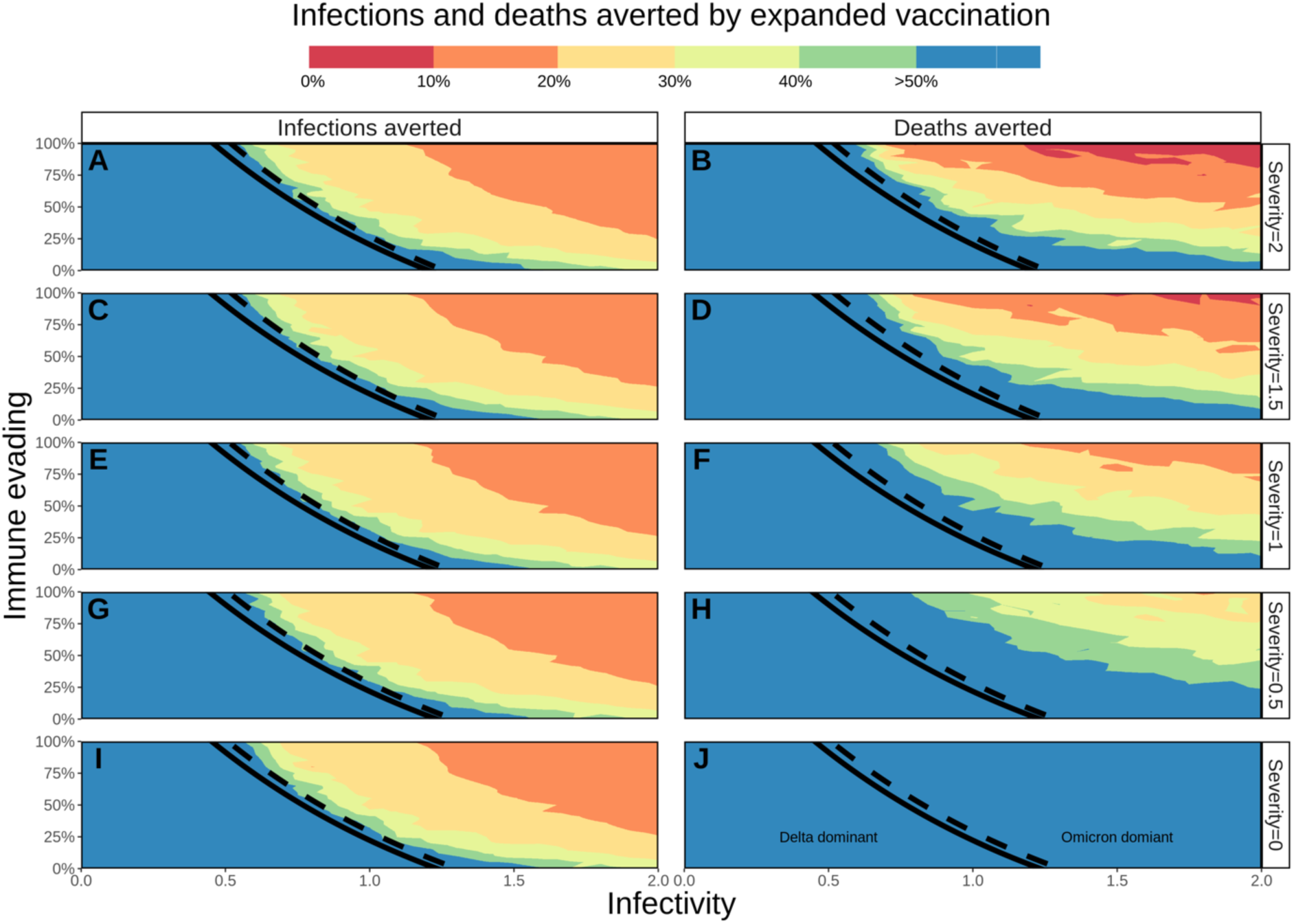
Percentage of COVID-19 infections and deaths averted by third-dose vaccines for adults and vaccinating 5-11-year-olds with doses one and two. The threshold (50% prevalence) for Omicron’s future dominance within 6 months is presented by black lines (solid line for expanded vaccination, dashed for no future vaccination). Left panels represent the percentage infections averted, right panels the percentage of deaths averted due to expanded vaccination, with colour representing % averted. Rows represent Omicron’s potential severity (0 to 2) relative to Delta (1), horizontal axes Omicron’s potential infectivity (0 to 2) relative to Delta (1) and left vertical axes Omicron’s potential immune evading capacity (0 to 100%). Maximum percentage of burden averted is capped at 50%, with blue area extending to values >50%.

**Fig. 3:**
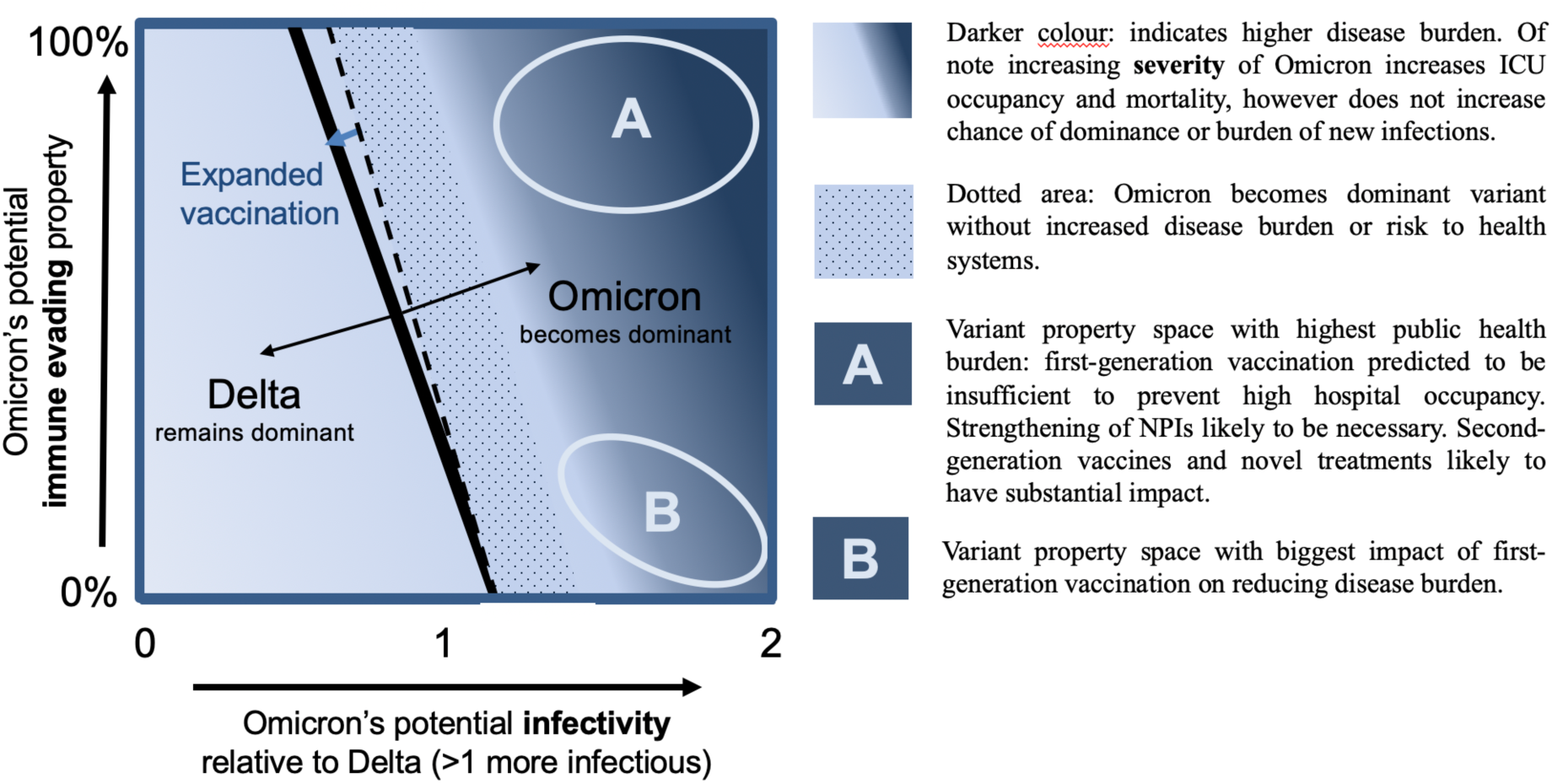
Schematic illustrating a summary of the interplay between potential infectivity, immune evasion of Omicron or other variant of concern on its chances of becoming dominant and increasing public health burden.

To aid interpretation of Omicron’s properties on its chance of becoming dominant and the associated overall impact on public health burden in a vaccinated population, we summarised our findings in a schematic (Fig. 3). The darkness of colour in the heat map indicates the level of disease burden (infections and deaths) and risk to health systems (hospital occupancy) at the corresponding level of relative infectivity and immune evasion. The dashed line indicates when Omicron would become the dominant variant without future vaccinations, the solid line when it would become dominant with expanded vaccination. The dotted area indicates when Omicron would become dominant without increased disease burden or risk to health systems. Area A indicates variant property space where expanded vaccination alone will not prevent increased transmission and would lead to increased infections, hospital occupancy, and deaths. Additional measures would be needed in the case of very high immune evading variant including second-generation vaccines and/or novel treatment once available, or strengthened NPIs in their absence. Area B indicates space where expanded vaccination with first-generation vaccines will have the highest impact on reducing disease burden and health system risk. In case of low severity in area B, expanded vaccinations may be sufficient for control.

## Discussion

Using our OpenCOVID individual-based transmission model, we simulated the interplay of Omicron’s potential combinations of infectivity, severity, and immune evasion properties. We first identified the threshold of Omicron becoming the dominant variant (over Delta), for which infectivity was found to be the main driver followed by immune evasion. Increased disease severity of Omicron was found to have limited effect on dominance, but when dominant, to allow for an increase in hospital occupancy and number of deaths. If Omicron has increased infectivity and severity to Delta, then expanding first-generation vaccination with a third-dose and vaccinating 5-11-year-olds could avert many cases and deaths. However, would Omicron be highly immune evading, first-generation vaccines would not suffice, and additional measures would be required to control transmission until vaccines are updated.

Besides additional preventive measures such as strengthened NPIs and second-generation vaccines, new treatments that have been approved in December 2021 can now additionally be implemented to reduce the public health burden. (19-25) Maximum hospital capacity will have to be considered as it differs between countries, ranging from 220 to 800 beds per 100,000-population capita in Europe. (18) In a worst-case scenario of highly immune evading and increased severity of Omicron, for many countries, our predictions reached or surpassed maximum hospital capacity. However, very high occupancy rates are unlikely to be reached as countries would implement additional measures sooner.

Although our analysis focuses on hospital occupancy and mortality as a higher priority public health risk, even rising SARS-CoV-2 infections due to lower severity Omicron may lead to a substantial risk of increased long-COVID (26) and should not be neglected in response planning. Furthermore, as we considered expanding vaccination with third-doses, the global inequity of vaccine access, and the selective pressure and emergence of new variants in settings with low vaccination rates is a global health emergency that needs to be addressed (27).

Our transmission model is based on assumptions that influence its outcomes. In our model predictions, we identified that Omicron’s potential severity had little effect on the number of new cases, also because no impact on viral load was assumed. Higher viral load in the model is associated with increased transmissibility. The two vaccination scenarios we present in this study, expanded vaccination (with a third-dose for those 12 years of age and older, and two-dose vaccines for children aged 5-11 years) and no future vaccination, are examples of two extreme scenarios. The reality will lie in between and will be region dependent. Waning immunity is assumed to reduce linearly to zero in approximately one year. However, should immunity wane much slower, then the population would be protected for a longer period against either Delta (or a new non-immune evading variant), and our outcomes would prove too pessimistic. Our outcomes would be too optimistic should immunity wane faster than assumed. A higher or lower value of 1.2 for the effective reproduction number at the start of the winter period, prior to introduction of Omicron, could have influenced peak hospital occupancy estimates, however the relative impact of vaccination is not sensitive to this parameter.

With Omicron being more infectious, less severe, and slightly immune evading relative to Delta, our model predicted that Omicron would indeed quickly become the dominant variant (within 1-4 months, depending on its exact combination of viral property values), leading to a peak in cases and hospitalisations (up to 50 hospital beds per 100,000 people with expanded vaccinations). For these scenarios we predicted that the expanded vaccination would prevent >40% of hospitalisations. In European countries with an expanded vaccination rollout, the hospital occupancy peak during the Omicron outbreak ranged between 20 and 50 beds per 100,000 population, echoing our simulations. (28)

SARS-Cov-2 mutations have been identified throughout the pandemic and new variants are likely to continue to emerge. Compared with the Alpha variant, Delta has severity ratios reported as 1.3- to 2.3-times higher for hospital admissions and deaths. (29) Both vaccine-acquired and naturally acquired immunity will not suffice to prevent infection or disease if a new variant is highly immune evading. Based on these predictions we stress that vaccines may need adjustment to respond to current or future viral mutations. We further advocate for continued genomic surveillance and see it crucial to quickly test and continue improving assays that elucidate immune evasion properties of a new VOC, to better understand the risk of increased infectivity, severity, and immune evasion.

## Conclusion

As the properties of Omicron and any future SARS-CoV-2 VOC become known, our analyses elucidate interpretation of the variant’s potential dominance and subsequent public health burden. Combined with VOC genomic (30) and population epidemiological surveillance (31), alongside antibody neutralisation testing (32) our findings provide crucial quantitative guidance to decision-makers at a critical time.

## Data Availability

Data sharing is not applicable to this article as no datasets were generated or analysed during the study. Data informing model parameters are described in Shattock et al. All model code and Figure code are open access and publicly available at (https://github.com/SwissTPH/OpenCOVID). For this paper model code and Figure code are version 2.0 of OpenCovid.

## Methods

### OpenCOVID individual-based model

OpenCOVID is a stochastic, discrete-time, individual-based transmission model of SARS-CoV-2 infection and COVID-19 disease (15). The model simulates viral transmission between infectious and susceptible individuals that come in contact through an age-structured, small-world network. The probability of transmission in each exposure is influenced by the infectiousness of the infected individual, the immunity of the susceptible individual (acquired through previous infection and/or vaccination), and a background seasonality pattern (reflecting a larger proportion of contacts being in closer contact indoors with cooler temperatures). Infectiousness is a function of viral variant infectivity and time since infection (which follows a gamma distribution peaking approximately at the time of symptom onset). Once infected, a latency period is followed by a pre-symptomatic stage, after which the individual can experience asymptomatic, mild, or severe infection. Severe cases can lead to hospitalisation, ICU admission, and ultimately death. Recovery after infection leads to development of immunity. This immunity is assumed to wane over time and can be further reduced if exposed to a novel variant with immune evading properties (Supplementary Table 2 for further details). The model has the capacity to represent a number of containment measures, including non-pharmaceutical interventions such as social distancing and facemask usage, testing strategies such as test-diagnose isolate, mass-testing, and contact tracing, and also pharmaceutical interventions such as vaccination and treatment.

Detailed model descriptions and model equations are described in. (15) Open access source-code for the OpenCOVID model is publicly available at (https://github.com/SwissTPH/OpenCOVID).

### Vaccination

In this analysis, we simulate the impact of mRNA vaccines Pfizer/BioNTech and Moderna, which together make up 78% of the total number of doses secured in Europe. (5) Fully susceptible, partially susceptible, and infected individuals not in hospital are considered eligible to receive a vaccine. Vaccines have a two-fold effect; first, they provide protection against new infection through development of immunity (90% reduced susceptibility). Secondly, once infected, vaccines reduce the probability of developing severe symptoms, leading to a 95% reduction in severe disease, impacting hospitalisations, ICU admissions, and deaths. We assume vaccination does not impact the probability of onward transmission once the individual is infected. Details regarding targeted vaccination groups and assumed durations between doses and vaccine efficacies are described in the Supplementary Information 2.2-2.3. Associated waning immunity profiles after infection and vaccination are described in Supplementary Information 2.3 and 2.4.

### Variant properties

Delta (B.1.617.2) is assumed to be the dominant transmission variant at the time of Omicron (B.1.1.529) emergence. A full factorial range of Omicron properties were considered:1) infectivity (transmission multiplication factor per exposure relative to Delta), ranging from 0 to 2, 2) disease severity (multiplication factor per infection, relative to Delta ranging from 0 to 2), and 3) immune evasion capacity from 0-100%. In the model, once infected with the new variant, severity influences the chance to manifest severe symptoms, rather than mild or no symptoms. One hundred percent immune evasion means fully evading any previously naturally or vaccination-acquired immunity, making an individual fully susceptible to infection with a new variant. The immune evasion property is assumed to only relate to the individual’s previously acquired immunity (their susceptibility), thus only impacting the rate of new infections and not to influence their severity of infection once infected. Full model details on the variant properties are described in Supplementary Information 2.5 alongside probabilities of immunologically naïve infected individuals developing severe disease in Supplementary Table 3.

### Model initialisation

All model simulations were designed to be pseudo-representative of a European setting at the beginning of December 2021. We assume 30% of the population have been previously infected with SARS-CoV-2 over a 630 day period (representing epidemic outbreak in Europe in March 2020). We assume the effective reproduction number (*R*_*e*_(*τ*)) on 1 December 2021, is equal to 1.2. This represents an average scenario of increasing case numbers across Europe at the start of the winter period prior to the emergence of Omicron. This level of *R*_*e*_(*τ*) is lower than levels in some European countries with strongly increasing cases as of early December, however also higher than those that implemented strong NPI before 1^st^ December. The average number of daily contacts required to achieve an *R*_*e*_(*τ*) of 1.2 inherently considers any non-pharmaceutical interventions in place at the beginning of the winter period in Europe prior to the emergence of Omicron. Seasonality is assumed to follow a cosine function, with a peak in seasonal infectivity occurring 6 weeks from model initialisation (representing mid-winter see Supplementary Information Figure 6).

### Analyses

For the full range of variant properties specified, we simulated two vaccination scenarios from the introduction of the new variant to six months into the future. For the first scenario, no future vaccinations are implemented. The second scenario is identical up to 1 December 2021, but simulates expanded vaccination with first-generation vaccines administering as third-doses in adults (six months after second-dose) and scale up of first- and second-dose in 5-17 year-olds. Each simulation provided the relative prevalence of Omicron over the next 6 months compared with Delta, as well as daily and cumulative numbers of new SARS-CoV-2 infections, maximum hospital occupancy, and COVID-19-related deaths. To reflect the element of chance that naturally occurs in transmission dynamics, 10 random stochastic simulations were performed per scenario for which we present the mean. In this analysis we do not explore the effect of varying NPI intensity over time. As such, our analysis reflects predicted disease dynamics and public health burden in the absence of strong NPIs, such as lockdowns.

## Data availability statement

Data sharing is not applicable to this article as no datasets were generated or analysed during the study. Data informing model parameters are described in Shattock *et al*. (15)

## Code availability statement

All model code and Figure code are open access and publicly available at (https://github.com/SwissTPH/OpenCOVID). For this paper model code and Figure code are version 2.0 of OpenCovid (at final submission a zendo DOI will be provided for the code and model generated data behind all figures).

## Acknowledgements

The authors want to acknowledge and thank the members of the Disease Modelling Unit, Swiss Tropical and Public Health Institute. Calculations were performed at sciCORE (http://scicore.unibas.ch/) scientific computing core facility at University of Basel

## Author contributions

EALR and AJS conceived the study, performed the analyses, prepared figures, and conducted model and analysis validation. AJS further developed the model with input from EALR, SLK, and MAP. All authors contributed to interpretation of the results, writing the draft, and final version of the manuscript, and gave final approval for publication.

## Competing interests

The authors declare no competing interests.

## Funding

Funding for the study was provided by the Botnar Research Centre for Child Health (to MAP) and the Swiss National Science Foundation Professorship of MAP (PP00P3_170702).

## Supplementary Information

### 1. Supplementary Figures 1-6 and Tables 1-3

**Supplementary Table 1.** Overview of two vaccination scenarios. No future vaccination, and expanded vaccination through third-dose in adults (six months after second-dose) and scale up in 5-17 year-olds with first-generation vaccines. Additional details are provided in section 2.3.

| Group | As of 1 December 2021 | No future vaccination | Expanded vaccination |
| --- | --- | --- | --- |
| **65+ or comorbidities** | 90% coverage | 90% coverage | 90% coverage of third dose* 180 days after second dose |
| **Adults 18-64** | 70% coverage | 70% coverage | 70% coverage of third dose* 180 days after second dose |
| **Adolescents 12-17** | 50% coverage | 50% coverage | Scale up to 70% coverage within 2 months |
| **Children 5-11** | 0% coverage | 0% coverage | Scale up to 50% coverage within 4 months |
| **Total number of doses in future** |  | zero | 16,000 per 100,000 people over six-months |

\*5% dropout included between those receiving second and third-dose (95% acceptance rate for third dose for these with two doses)

**Supplementary Table 2.** Summary of seven combinations of Omicron’s or any future variant of concern’s potential properties including infectivity, severity, and immune evasion. Probability of the new variant to become the dominant variant, associated public health burden, and the effect of expanded vaccination through third-dose in adults (six months after second-dose) and scale up in 5-17 year-olds with first-generation vaccines.

| Simulation | Infectivity (relative to Delta) | Severity (relative to Delta) | Immune evasion | Probability of Omicron dominance | Public health impact | Expanded vaccination* effect |
| --- | --- | --- | --- | --- | --- | --- |
| **1** | 1 | 1 | 0% | Low | Negligible | High |
| **2** | 2 | 1 | 0% | Very high | High | High |
| **3** | 2 | 2 | 0% | Very high | High | Very high |
| **4** | 1 | 1 | 100% | Very high | High | Moderate |
| **5** | 1.2 | 2 | 20% | High | Low | Moderate |
| **6** | 1.5 | 2 | 50% | Very high | High | Low |
| **7** | 2 | 2 | 100% | Very high | Very high | Negligible |

\* Expanded vaccination through third-dose in adults and vaccinating 5-17 year-olds with first-generation vaccines

**Supplementary Figure 1.**
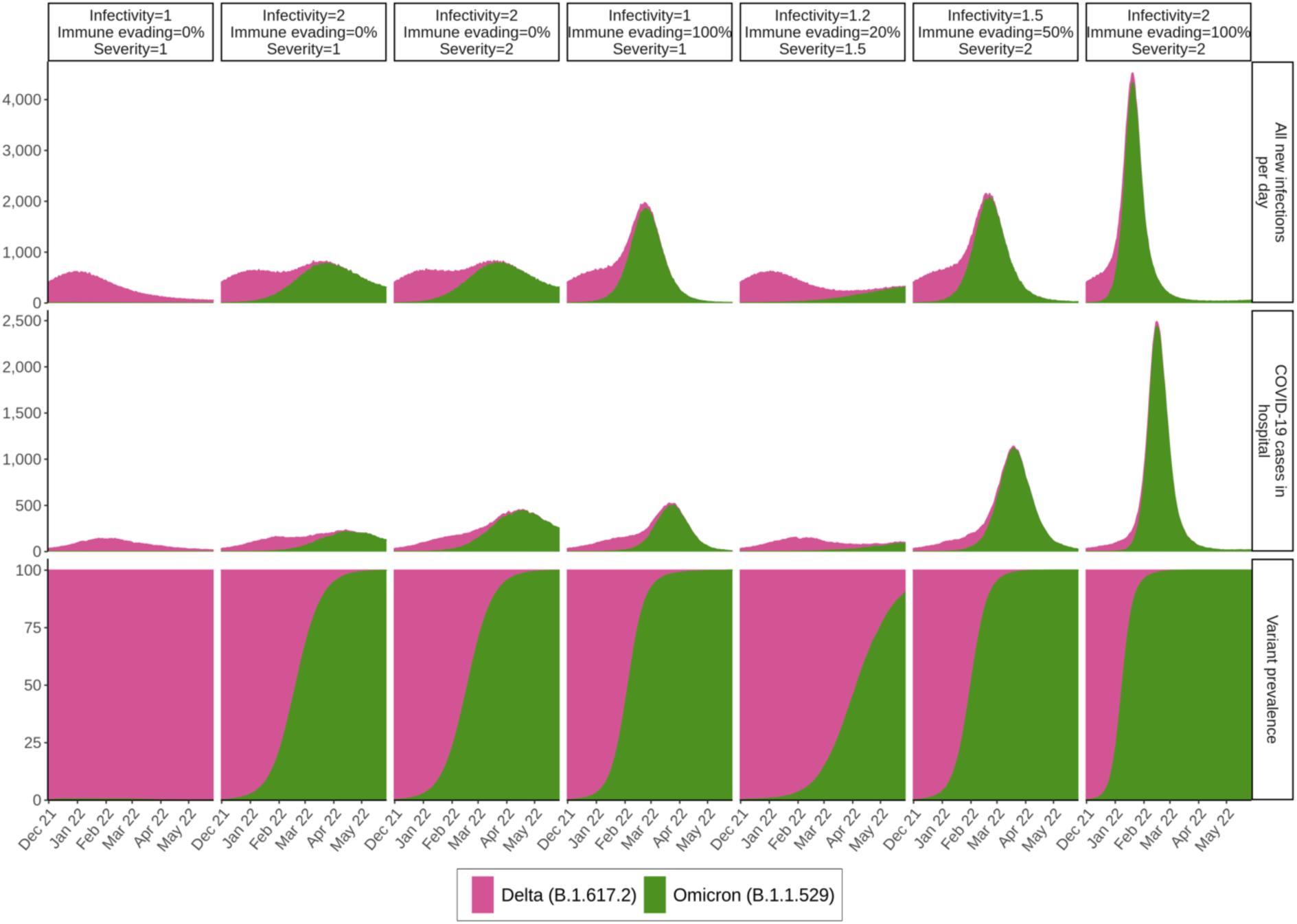
Temporal epidemiological trends for seven combinations of the new variant’s potential properties under no future vaccination. Public health burden is represented by COVID-19 infections and hospital occupancy per 100,000 population per day. Variant prevalence percentage over time is presented in the bottom row.

**Supplementary Figure 2.**
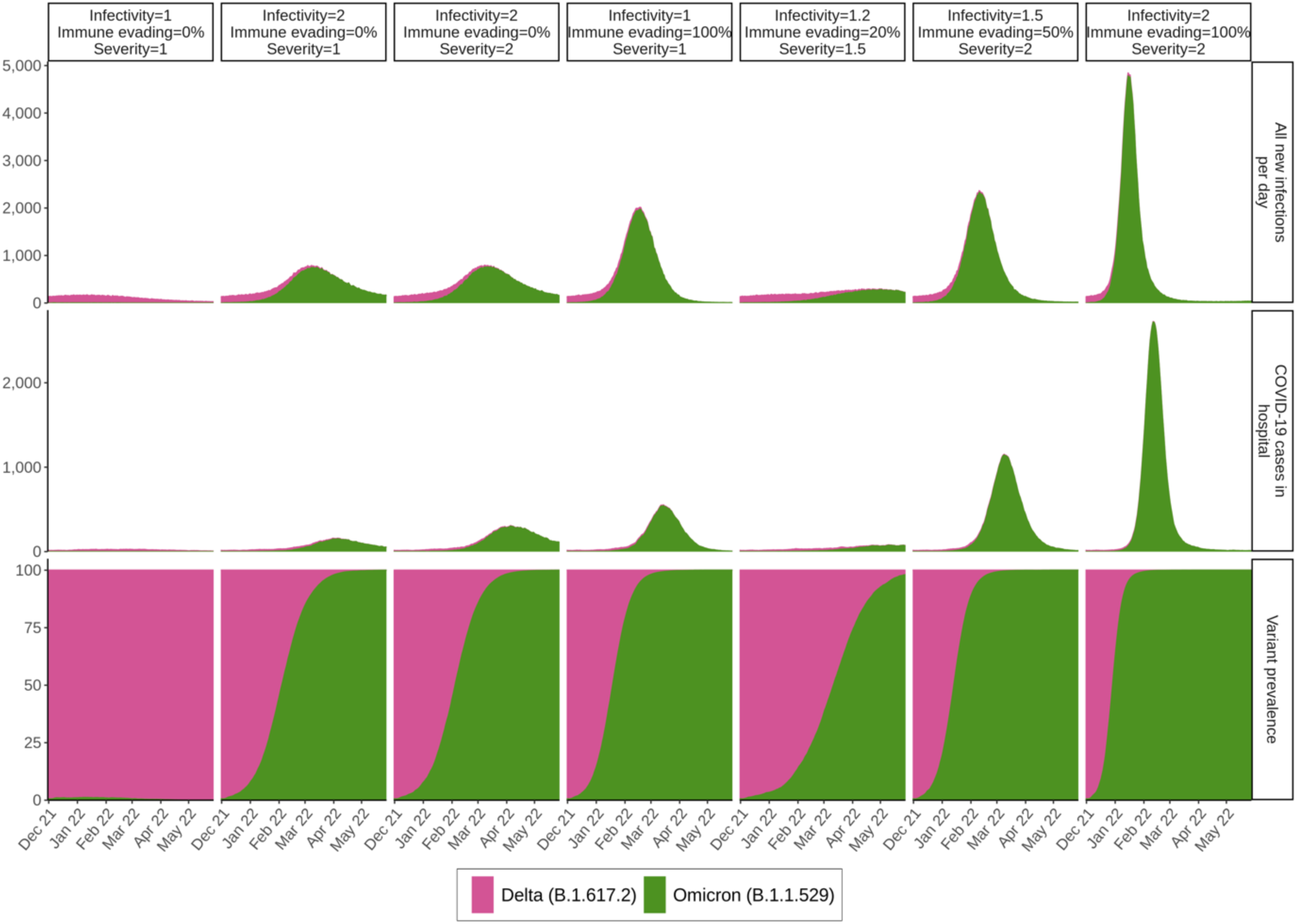
Temporal epidemiological trends for seven different combinations of Omicron’s potential properties under expanded vaccination. This figure represents a setting with future expanded vaccination through third-dose in adults (six months after second-dose) and scale up in 5-11 year-olds with first-generation vaccines. Public health burden is represented by COVID-19 infections and hospital occupancy per 100,000 population per day. Variant prevalence percentage over time is presented in the bottom row.

**Supplementary Figure 3.**
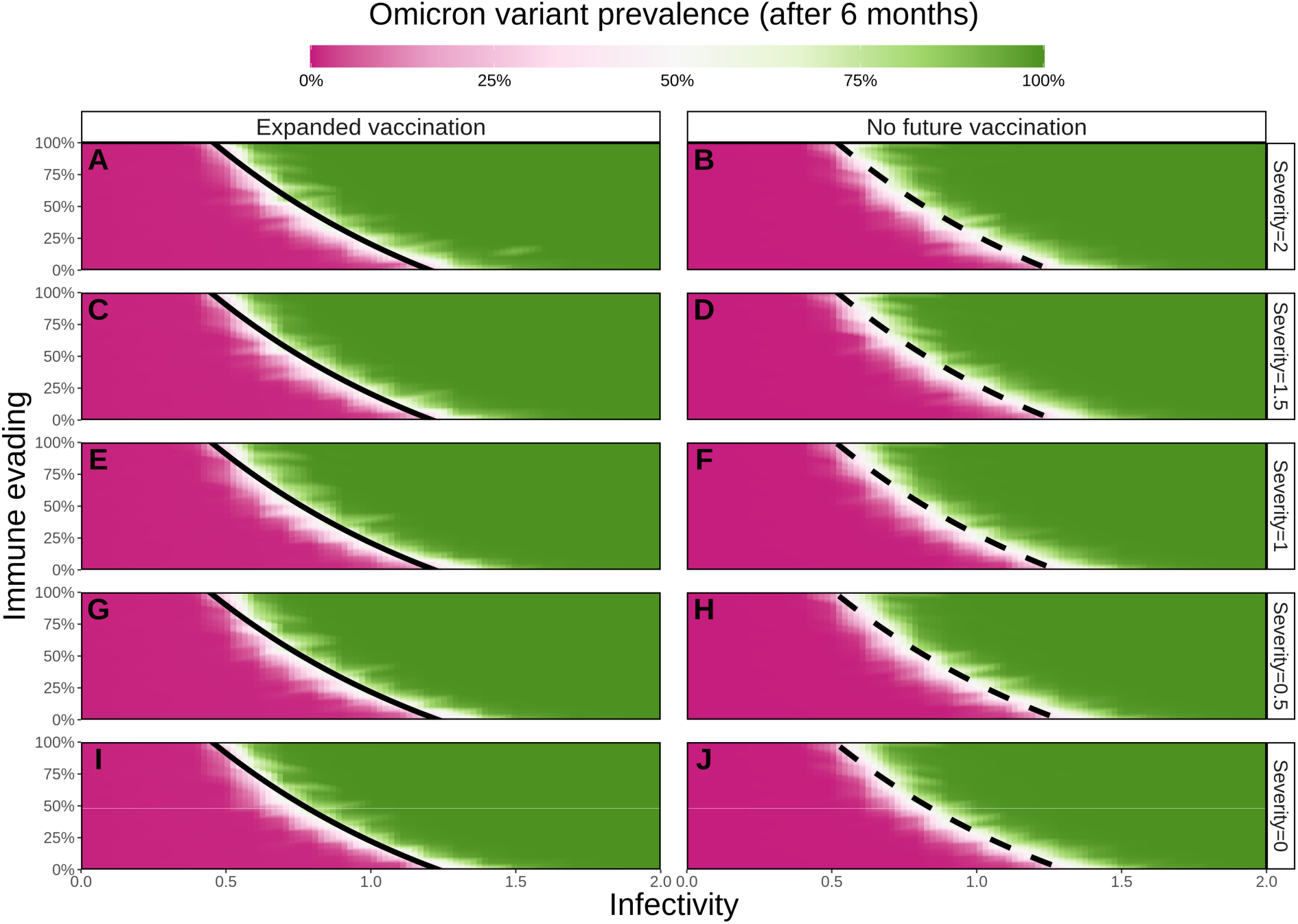
The projected prevalence of the new variant under combinations of infectivity, immune evasion capacity, and severity after six months. Black line represents the threshold (50%) for the new variant to become dominant (areas to the right of the lines). Left column represents a setting with expanded vaccination through third-dose in adults (six months after second-dose) and scale up in 5-17 year-olds with first-generation vaccines. Right column represents a setting with no future vaccination. Horizontal axes represent the range of the new variant’s potential infectivity (0 to 2) relative to Delta (1). Vertical axes represent the range of the new variant’s potential immune evasion capacity (0 to 100%). Rows represent five levels of the new variant’s potential severity (0 to 2) relative to Delta (1).

**Supplementary Figure 4.**
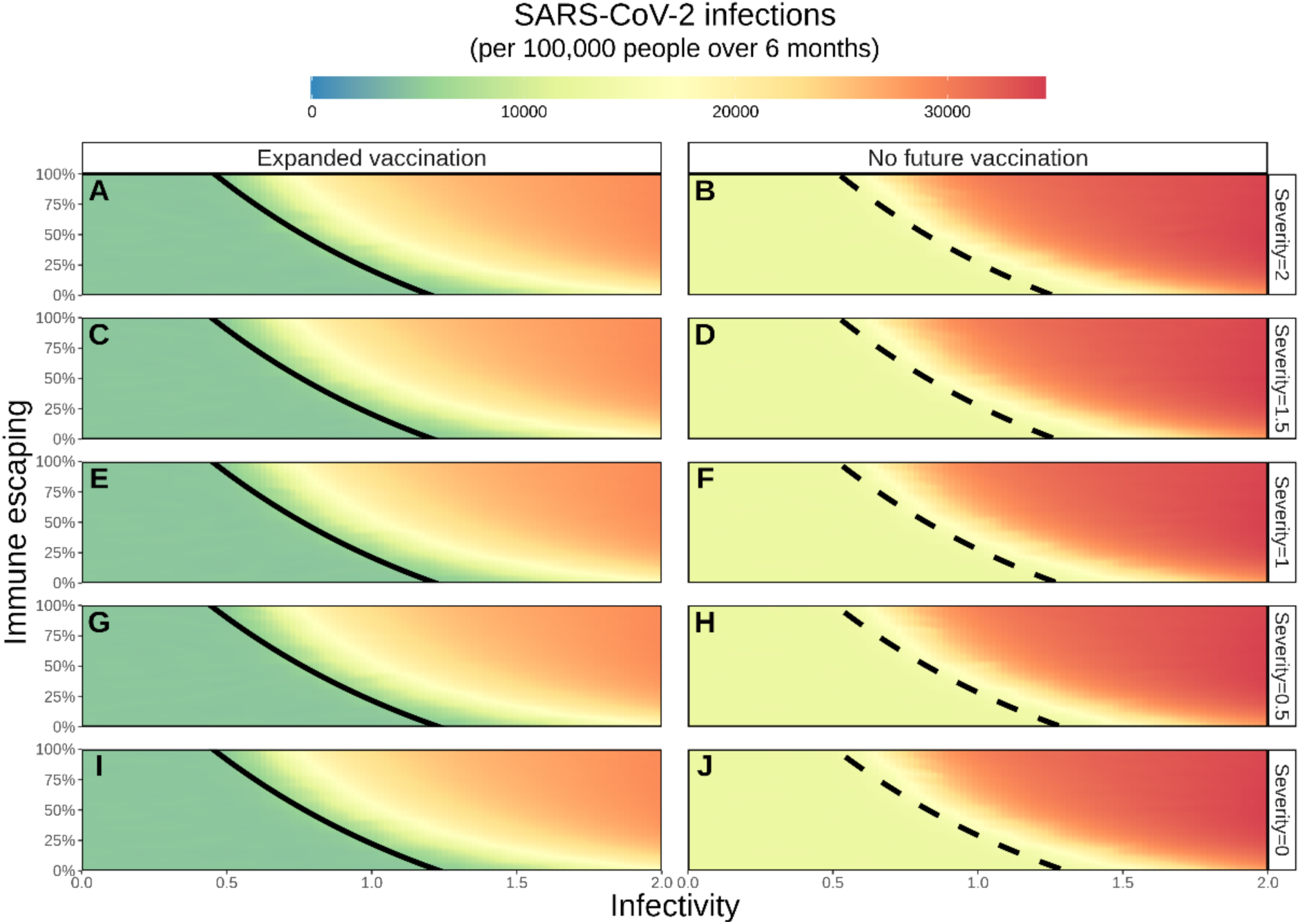
Cumulative number of SARS-CoV-2 infections (per 100,000 population over the six-month simulation period) for a wide range of variant properties. Solid and dashed black lines represent the threshold (50%) for the new variant to become dominant (area to the right of the black line). Left panels represent a setting with expanded vaccination with first-generation vaccines through third-dose in adults (six months after second-dose) and scale up in 5-17 year-olds, right panels, a setting with no future vaccination. Horizontal axes represent the range of the new variant’s potential infectivity (0 to 2) relative to Delta (1). Left vertical axes represent the range of the new variant’s potential immune evasion capacity (0 to 100%). Rows represent five levels of the new variant’s potential severity (0 to 2) relative to Delta (1).

**Supplementary Figure 5.**
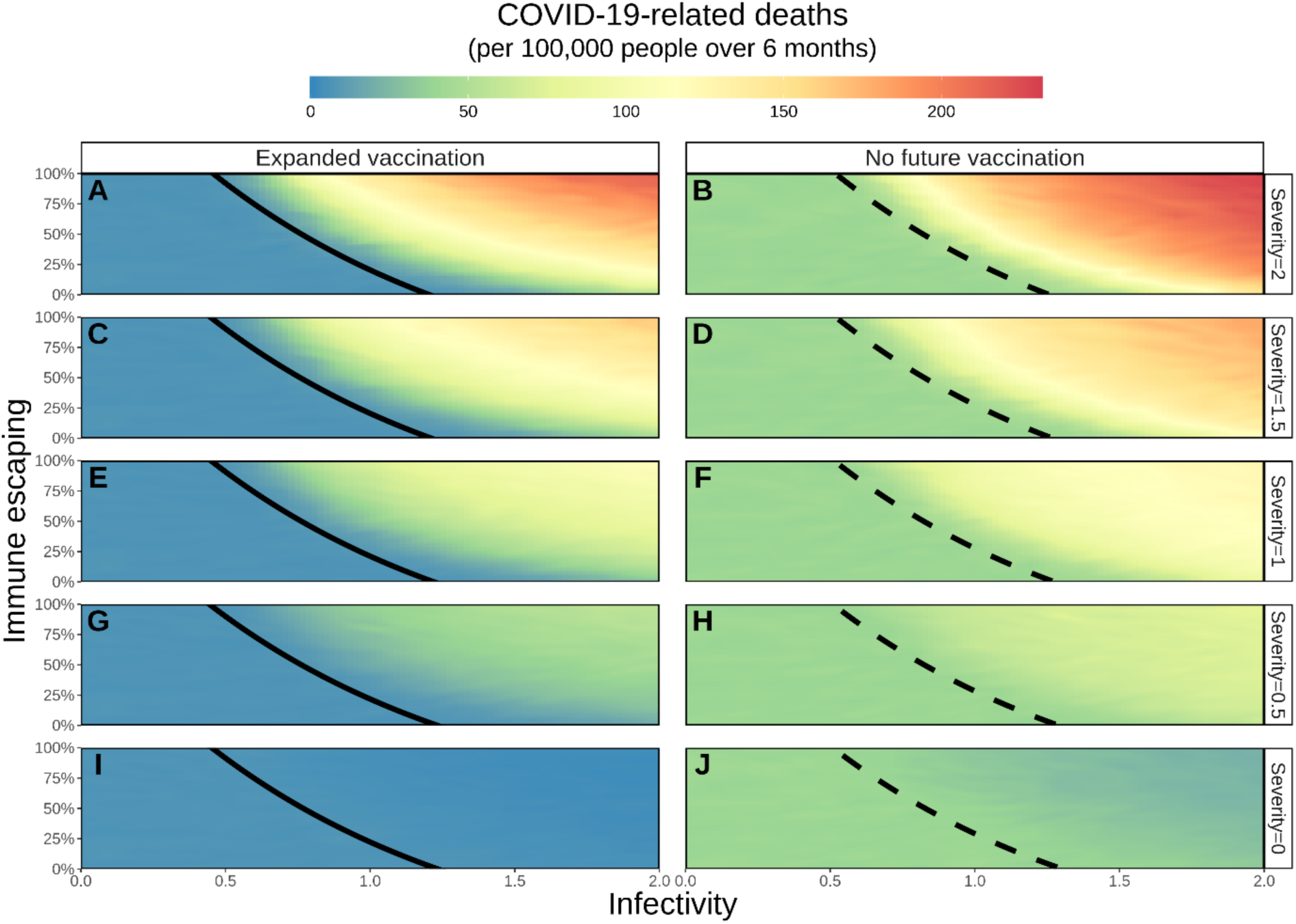
Cumulative number of COVID-19-related deaths (per 100,000 population over the six-month simulation period) for a wide range of variant properties. Solid and dashed black lines represent the threshold (50%) for the new variant to become dominant (area to the right of the black line). Left panels represent a setting with expanded vaccination through third-dose in adults (six months after second-dose) and scale up in 5-17 year-olds using first-generation vaccines, right panels, a setting with no future vaccination. Horizontal axes represent the range of the new variant’s potential infectivity (0 to 2) relative to Delta (1). Left vertical axes represent the range of the new variant’s potential immune evasion capacity (0 to 100%). Rows represent five levels of the new variant’s potential severity (0 to 2) relative to Delta (1).

**Supplementary Figure 6.**
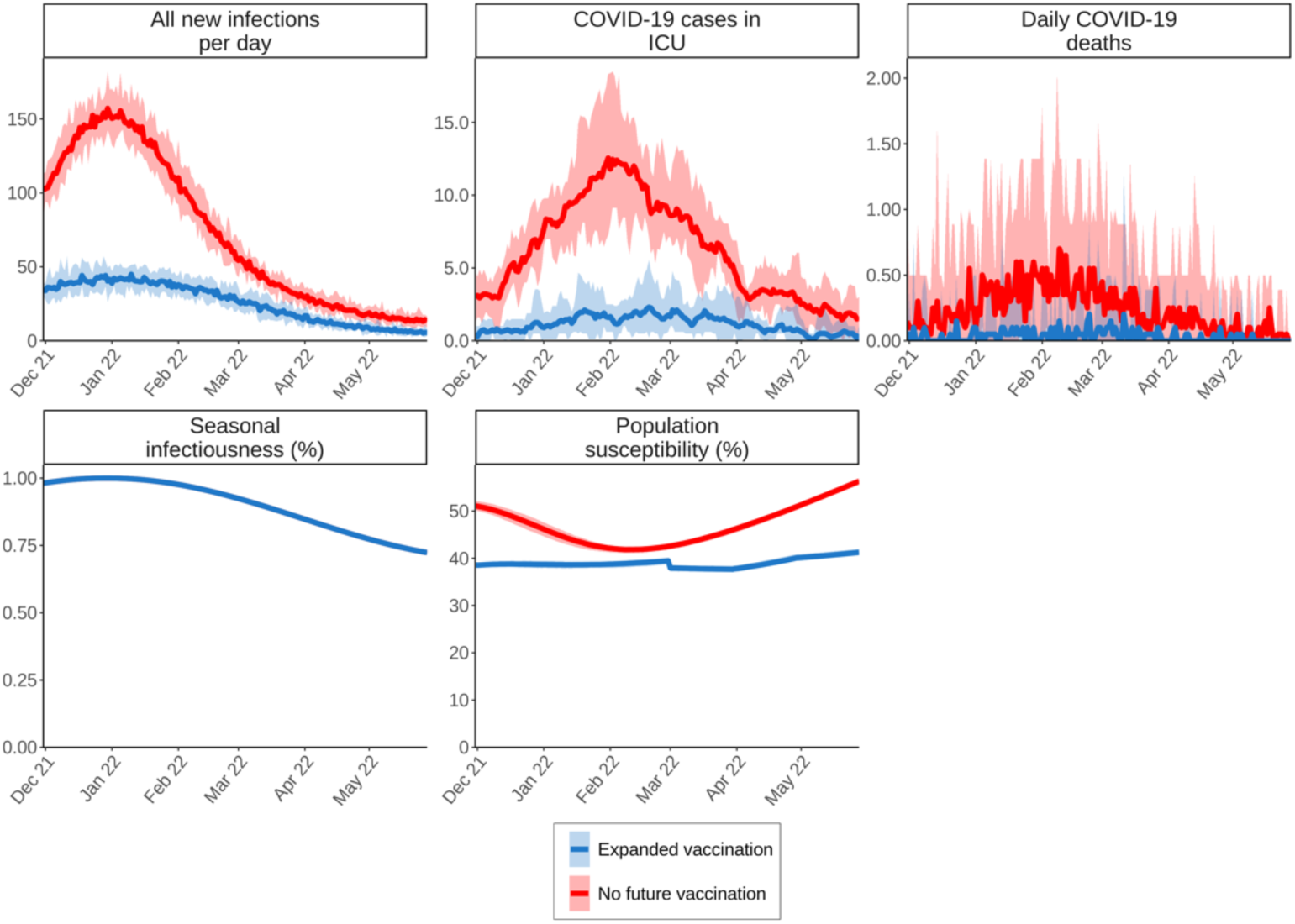
Predicted temporal epidemiological trends for Delta in the absence of Omicron under no future vaccination and expanded vaccination, alongside seasonality profile. Public health burden is represented by COVID-19 infections, ICU occupancy and COVID-19 related mortality per 100,000 population per day. Seasonal infectious and susceptible population over time is presented in the bottom row. Blue illustrates dynamics with expanded vaccination, and red with no future vaccination.

### 2. Supplementary methods

#### 2.1 Model initialisation

All model simulations were designed to be pseudo-representative of a general Western European setting at the beginning of December 2021. We assume 30% of the population have been previously infected with SARS-CoV-2 over a 630 day period (representing epidemic outbreak in Europe in March 2020). The fraction that was both previously infected and vaccinated prior to the emergence of the Omicron variant was age-dependent and ranged from 3% in 10-20-year-olds to 30% in 80-90-year-olds (see section 2.2 for further details on vaccination rates prior to the Omicron’s emergence). We assume the effective reproduction number on 1 December 2021 is equal to 1.2. This represents increasing case numbers across Europe at the start of the winter period, prior to the emergence of Omicron. The average number of daily contacts required to achieve an initial effective reproduction number of 1.2 inherently considers any non-pharmaceutical interventions in place at the beginning of the winter period in Europe prior to the emergence of Omicron. Seasonality is assumed to follow a cosine function, with a peak in seasonal infectivity occurring 6 weeks from model initialisation (representing mid-winter, Supplementary Figure 6).

#### 2.2 Vaccine rollout

Four distinct risk groups are simulated for vaccination rollout; those 65 years of age and older or live with comorbidities (high-risk group), all other adults (18-64-year-olds), adolescents (12-17-year-olds), and children (5-11-years-olds). In our simulations, vaccinations start in the high-risk group in January 2021 reaching a 90% coverage rate on 1 December 2021. The 18-64-year-olds start vaccination on 1 May 2021, achieving 70% coverage on 1 December 2021. Adolescents 12-17 years of age started vaccinations on 1 October 2021 with a coverage of 50% achieved by 1 December 2021. Children 5-11 years of age start vaccinations of doses 1 and 2 on 1 December 2021 and are only included as part of the extended vaccination scenario. Vaccination groups, associated vaccination coverages as of 1 December 2021, and the simulated future scenarios are summarized in Supplementary Table 1.

#### 2.3 Vaccine-induced immunity profile

We model an interval of 28 days between the first and second dose, and a maximum of 95% vaccine efficacy to be reached 14 days after the second dose (increasing with a sigmoidal curve). We assume that 90% of vaccination effect is transmission blocking (i.e. a 90% reduced probability to become infected after being exposed). We assume vaccinated individuals will always be administered two doses and assume that 95% of those vaccinated with doses one and two will accept a third-dose. Third-doses are administered 6 months after the second dose, after which the maximum vaccine efficacy of 95% is again reached 14 days after the last dose has been administered. One month after the last dose, immunity starts waning linearly to zero over 335 days ^1,2^. The left panel of Supplementary Figure 7 provides a schematic overview of the waning immunity pattern after vaccination.

#### 2.4 Infection-induced immunity profile

After recovering from infection with SARS-CoV-2, individuals develop naturally acquired immunity with a maximum level of 90% reduced susceptibility which they maintain for a month, after which their immunity wanes linearly to zero over a period of 335 days ^1,2^. The level of immunity is the reverse of the susceptibility of the individual, which thus increases over time. The Delta variant is assumed not to be immune evading, therefore immunity following infection from Delta and Omicron provides a similar risk of infection when exposed to Delta. The right panel of Supplementary Figure 7 provides a schematic overview of the waning immunity pattern after natural infection.

**Supplementary Figure 7.**
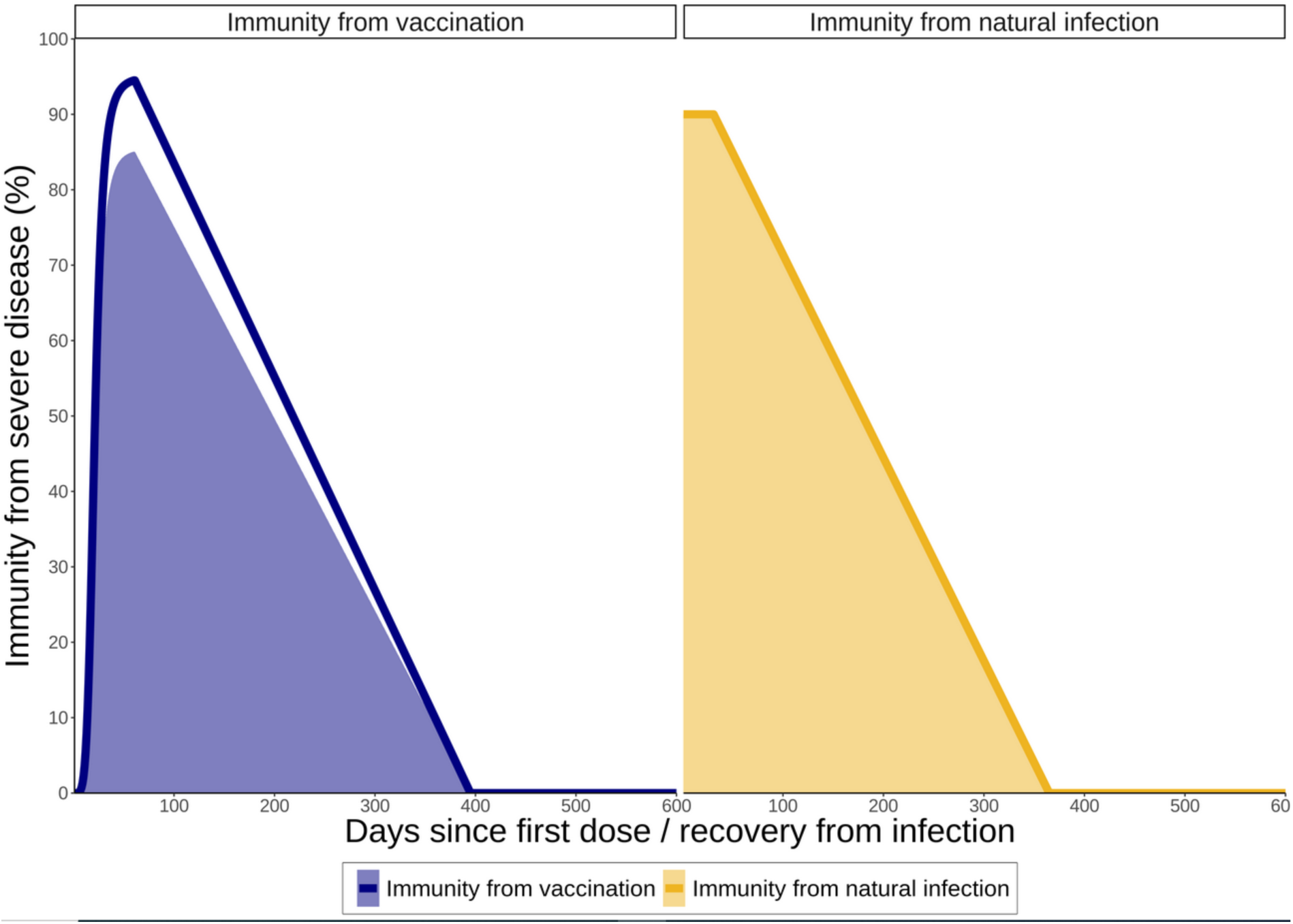
Profiles for immunity acquired from a) vaccination, and b) natural infection. The shaded areas represent immunity from transmission when exposed to a variant without immune escaping properties. The solid line represents overall immunity to severe disease.

#### 2.5 Effect of variant properties and vaccination on prognosis

The individual’s prognosis depends on multiple factors including age, vaccine status, co-morbidity, and variant severity. The probabilities of 1) a symptomatic case developing severe disease, 2) a severe case becoming critical, and 3) a critical case ultimately leading to death, are all defined as functions for the above-mentioned factors. For this study, we use probabilities reported in ^3^, updated to represent the additional risk of hospitalisation from infection with VOC Delta (B.1.617.2) ^4-6^. In additional to age-related risk, the probability that an infected individual will develop severe symptoms is also scaled by the severity factor of the viral variant exposed to.

In this study we use a severity factor of 1 for VOC Delta (B.1.617.2), and consider a range of potential relative severity factors for VOC Omicron (B.1.1.529) between 0 and 2. That is, a variant that has 0%-200% severity of Delta. For vaccinated individuals that become infected (noting that the transmission-blocking action of the vaccine reduces the probability of infection), the probability of developing severe disease is reduced by the severity or disease-blocking property of the vaccine. The level to which the probability of severe disease is reduced is dependent upon the level of immunity at the time of infection.

Vaccine-induced immunity is assumed to wane over time and can be further decreased if exposed to a variant with immune evading capacity. In this study, we consider the full range of potential immune evading properties of Omicron, from 0% to 100%. The probability that immunologically naïve (i.e., unvaccinated, and previously uninfected) individuals with no comorbidities infected with Delta (severity factor 1, 0% immune evading) develop severe disease are given in Supplementary Table 3.

**Supplementary Table 3.** Probabilities that immunologically naïve (i.e., unvaccinated and previously uninfected) individuals with no comorbidities infected with Delta (severity factor 1, 0% immune evading) develop severe disease (source ^3-6^)

| Age group | Severe disease<br>(Delta, unvaccinated) |
| --- | --- |
| **0-10** | <1.0% |
| **10-20** | 1.0% |
| **20-30** | 1.2% |
| **30-40** | 2.4% |
| **40-50** | 3.9% |
| **50-60** | 6.6% |
| **60-70** | 16.3% |
| **70-80** | 26.0% |
| **80-90+** | 33.0% |

## Notes

### Competing Interest Statement

The authors have declared no competing interest.

### Summary of Updates

We updated several figures and text related.

